# Expansion of *tetM*-carrying *Neisseria gonorrhoeae* in the US, 2018-2024

**DOI:** 10.1101/2025.03.24.25324420

**Authors:** David Helekal, Tatum D. Mortimer, Yonatan H. Grad

**Affiliations:** Department of Immunology and Infectious Diseases, Harvard T. H. Chan School of Public Health, Boston, Massachusetts, USA; Department of Population Health, College of Veterinary Medicine, University of Georgia, Athens, Georgia, USA

## Abstract

Doxycycline use for bacterial sexually transmitted infections (STIs) increased in recent years due to changes in treatment guidelines, the shortage of benzathine penicillin for treatment of syphilis, and adoption of doxycycline post-exposure prophylaxis. While this increased use is expected to select for doxycycline resistance, particularly in *Neisseria gonorrhoeae*, the impact has been unclear. Here, we analyzed over 14,000 publicly available *N. gonorrhoeae* genome sequences from 2018-2024 generated through the US Centers for Disease Control and Prevention *N. gonorrhoeae* surveillance system, assessed the distribution of *tetM*, a plasmid-borne gene that confers high-level (MIC ≥ 16µg/mL) tetracycline resistance, and evaluated the spatial, temporal, and evolutionary dynamics of its spread. The proportion of *Neisseria gonorrhoeae* carrying *tetM* increased from below 10% in 2020 to over 35% in 2024.

Phylogenetic analysis revealed four major clades that have rapidly expanded. Two of these lineages also carry *penA* alleles that increase resistance to ceftriaxone. The strength of selection for tetracycline resistance, as indicated by its increased proportion and the growth of major *tetM*-carrying lineages, suggests a favorable environment for *N. gonorrhoeae* strains spreading globally that carry *tetM* and resistance to multiple other antibiotics used in gonorrhea treatment.

## MAIN TEXT

Doxycycline use for bacterial sexually transmitted infections (STIs) increased in recent years due to changes in treatment guidelines, the shortage of benzathine penicillin for treatment of syphilis, and adoption of doxycycline post-exposure prophylaxis (doxy-PEP). While this increased use is expected to select for doxycycline resistance, particularly in *Neisseria gonorrhoeae*, the impact has been unclear. Here, we analyzed over 14,000 publicly available *N. gonorrhoeae* genome sequences from 2018-2024 generated through the US Centers for Disease Control and Prevention *N. gonorrhoeae* surveillance system, assessed the distribution of *tetM*, a plasmid-borne gene that confers high-level (MIC ≥ 16µg/mL) tetracycline resistance, and evaluated the spatial, temporal, and evolutionary dynamics of its spread (Supplementary Appendix).

The mean estimated proportion of isolates in this collection carrying *tetM* increased from below 10% in 2020 to >30% in the first quarter of 2024 (Figure 1A), with the highest prevalence in Health and Human Services Region 10 (Supplementary Appendix), consistent with a recent report from Seattle^1^. The increase in *tetM* prevalence in 2020 coincides with the shift away from azithromycin and towards doxycycline for treatment of chlamydia^2^, and the increase starting in the summer of 2022 coincides with the reporting of the DoxyPEP trial’s results^3^.

**Figure 1.**
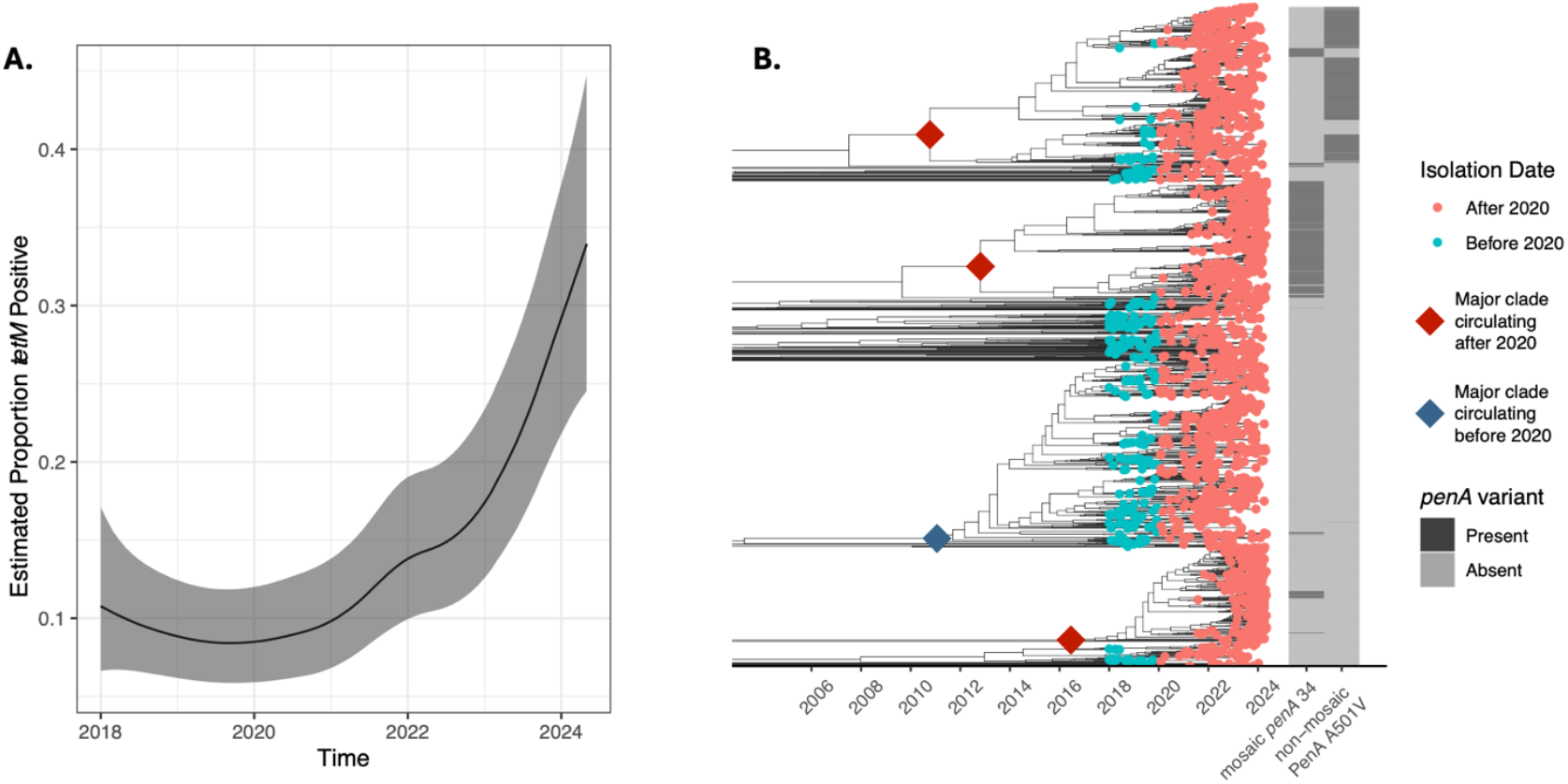
Expansion of *N. gonorrhoeae* carrying the *tetM* resistance determinant. A. The estimated proportion of isolates carrying *tetM* in a national surveillance dataset of *N. gonorrhoeae* from 2018 through the first quarter of 2024. The black line represents the modeled median estimate from data from HHS regions 2-10 (no data were available for HHS region 1) and the grey area represents the 95% credible interval. B. The phylogenetic relationships among *tetM*-carrying *N. gonorrhoeae* isolates reported in the dataset (see Supplementary Appendix for the full phylogeny with the deep branches and isolates without *tetM*). The diamonds indicate the roots of the lineages of *tetM* carrying isolates, with blue indicating the major *tetM* lineage circulating in the US before 2020 and red the major lineages after 2020. The circles indicate *N. gonorrhoeae* specimens; blue denotes those isolated before 2020 and red those isolated after 2020. The annotation columns to the right of the phylogeny indicate the presence or absence of *penA* variants, mosaic *penA* 34 and the A501V mutation, that increase resistance to cephalosporins.

The number of large *tetM*-carrying lineages increased from one in 2018-19 to four after 2020 (Figure 1B). Each of these four lineages rapidly expanded over the study period (Figure 1B). The major *tetM* lineage before 2020 accounted for 41% of *tetM-*carrying isolates, with the remainder sporadic isolates and small lineages. In contrast, the four major *tetM* lineages after 2020 together account for 83% of *tetM*-carrying isolates, indicating substantial expansion of these lineages. Of the four major lineages, two carry variants in *penA*, which encodes PBP2, the main target of ceftriaxone, that increase ceftriaxone resistance: A501V and mosaic *penA* 34 (Figure 1B; Supplementary Appendix)^4^. These results suggest that widespread use of doxycycline has rapidly reshaped the gonococcal population in the US.

Extremely drug-resistant *N. gonorrhoeae* lineages carrying both *tetM* and *penA* alleles such as mosaic *penA* 60 that is associated with a ceftriaxone MIC of 0.5-1 µg/mL, as well as resistance to fluoroquinolones and macrolides, have been spreading globally^5^. The strength of selection in the US for *tetM*, as indicated by its increased proportion and the number and growth of major *tetM*-carrying lineages, suggests a favorable environment for these highly drug-resistant strains to spread within the US.

## Data Availability

The data required to reproduce this analysis is publicly available through the NCBI Pathogen Detection database with detailed instructions available in the Supplementary Appendix.

## Acknowledgements/Funding

This work was supported by NIH R01 AI132606 and R01 AI153521 to Y.H.G. We thank the CDC and CDC-funded public health programs for their work collecting and characterizing *Neisseria gonorrhoeae* isolates and for making resulting *N. gonorrhoeae* genome sequences publicly available at NCBI.

