## Supplementary Appendix for "Expansion of *tetM*-carrying *Neisseria gonorrhoeae* in the US, 2018-2024"

### Methods

#### Data Acquisition

We searched the NCBI Pathogen Detection database (https://www.ncbi.nlm.nih.gov/pathogens/) for *Neisseria gonorrhoeae* isolate genome sequence and metadata collected by the US Centers for Disease Control and Prevention (CDC) with a collection date not before January 1, 2020 and a create date not after December 16, 2024 [1], which corresponded to including samples collected up to May 2024 . We filtered out isolates that were not identified as a *GCWGS* isolate, the designation given to isolates collected in the context of CDC *N. gonorrhoeae* surveillance. We included published CDC surveillance isolates from years 2018-2019 [2, 3]. These sets of isolates were collected as a part of CDC GISP [4], eGISP [4], and SURRG [5] programs. Given the sampling schemes, the representativeness of the collections cannot be guaranteed.

#### Genomic Analysis

*De novo* assembly was performed using SPAdes v 3.12.0 [6] with the –careful flag, and reference-based mapping to NCCP11945 (NC_011035.1) was done using BWA-MEM v 0.7.17 [7]. We used Pilon v 1.23 to call variants (minimum mapping quality: 20, minimum coverage: 10X) [8] after marking duplicate reads with Picard v 2.20.1 (https://broadinstitute.github.io/picard/) and sorting reads with samtools v 1.17 [9]. We generated pseudogenomes by incorporating variants supported by at least 90 % of reads and sites with ambiguous alleles into the reference genome sequence. We mapped reads to a single copy of the locus encoding the 23S rRNA and called variants using the same procedures [10].

For seven isolates we were unable to obtain a valid assembly after 3-days of run time.

Single nucleotide variants in *penA* were identified from pseudogenomes based on variant calls after mapping to the NCCP11945 reference genome. To determine the presence or absence of *tetM* and mosaic *penA* alleles, we used the results of blastn v 2.9.0 [11] searches of *de novo* assemblies for *tetM* (MG874353.1) and *penA* (from FA1090, NC_002946.2). We determined that *tetM* was present if the blast hit was >50% of the query length with >95% identity. We typed mosaic *penA* alleles according to the nomenclature in the NG-STAR database [12].

We filtered assembled genomes based on the following criteria: (1) The total assembly length was longer than 1900000 bp and less than 2300000 bp. (2) Reference coverage was more than 30%. (3) Percentage of reads mapped to reference was at least 70%. (4) Less than 12% of positions were missing in pseudogenomes. This resulted in (n = 14400) retained samples.

#### Phylogenetic Reconstruction

We used GUBBINS [13] to estimate recombining regions and IQTREE [14] for phylogenetic reconstruction. The sequence evolution model was *GTR+G+ASC* as selected by using MODELFINDER [15]. We ran Gubbins for 20 iterations. For the phylogenetic tree of all samples carrying *tetM*, we retained all sequences in which *tetM* was identified as present, resulting in ($n = 2331$) sequences. For the phylogenetic tree representative of the entire dataset, we first down-sampled the sequences so that within each calendar year quarter no more than $n = 250$ sequences were retained. If there were fewer than 250 sequences in a given quarter, all sequences for that quarter were retained. If there were more than 250 sequences in a given quarter, a sample of 250 sequences was drawn uniformly without replacement from all sequences in that given quarter. This resulted in a total of ($n = 6460$) sequences.

We estimated the timing of ancestral nodes and molecular clock parameters of the tree reconstructed by GUBBINS with BACTDATING [16] under the additive relaxed clock model [17].

Trees were annotated and visualized using the package GGTREE [18].

#### *tetM* Frequency Analysis

To obtain pooled estimates of *tetM* frequency trajectories smoothed over time and HHS regions, we first filtered the dataset to remove all entries that had a missing value in one of the following fields: (1) location; (2) collection date; (3) presence of *tetM*. We excluded isolates where the location field could not be unambiguously parsed as one of the ten HHS regions, enumerated as $1 - 10$. As there was no data for HHS region 1 for the years spanning 2020-2024, this region was excluded from further analysis.

To analyze the overall smoothed *tetM* trend as well as the smoothed *tetM* trajectories per HHS region, we used INLA version `24.04.25-1` to fit a Bayesian hierarchical panel timeseries model [19]. For each month indexed by $t$ and in each HHS region indexed by $i$, we considered the number of *tetM*-positive samples $Y_{i,t}$ out of the total number of samples in that given month and region $N_{i,t}$ as binomially distributed

$$Y_{i,t} \sim \texttt{binom}(\sigma^{-1}(y_{i,t}), N_{i,t}) \tag{S1}$$

where $\sigma^{-1}(\cdot)$ denotes the inverse of the logit link-function and $y_{i,t}$ is the linear predictor for region $i$ in month $t$.

We modeled the logit of the frequency $y_{i,t}$ in HHS region $i$ at month $t$ as a combination of of a global trend and HHS region specific random effects. The HHS region specific random effects consisted of (1) AR1-distributed fluctuations around the global trend and (2) a constant offset term. This model can be described by the following regression equation:

$$y_{i,t} = \mu \mid i + f_2(t|i) + f_1(t) \tag{S2}$$

The random per-HHS region intercept is denoted by $\mu \mid i$ and is assumed to be identically and independently distributed according to a Gaussian distribution with a 0 mean and unknown precision:

$$\mu \mid i \sim \mathcal{N}(0, \tau_{mu}^{-1}) \quad i.i.d. \tag{S3}$$

For each HHS-region, the random fluctuations over time are are denoted by $f_2(t|i,)$ and are assumed to be identically and independently distributed between regions according to an AR1 process with unknown parameters $\tau_{hhs}$ and $\rho$:

$$\begin{aligned} f_2(t+1|i) - \rho f_2(t|i) &\sim \mathcal{N}(0, \tau_{hhs}^{-1}) \quad i.i.d. \\ f_2(1) &\sim \mathcal{N}(0, (\tau_{time}(1-\rho^2))^{-1}) \end{aligned} \tag{S4}$$

Finally, $f_1(t)$ denotes the global trend modeled as a second order random walk RW2 with unknown precision $\tau_{time}$:

$$f_1(t+2) - 2f_1(t+1) + 2f_1(t) \sim \mathcal{N}(0, \tau_{time}^{-1}) \quad i.i.d. \tag{S5}$$

For all precision parameters, we used a half-normal prior imposed on the corresponding inverse-precision parameters:

$$\tau_{time}^{-1}, \tau_{hhs}^{-1}, \tau_{mu}^{-1} \sim \mathcal{N}(0,1)\mathbb{1}_{[0,\infty)} \quad i.i.d. \tag{S6}$$

For the AR1 parameter $\rho$, we used the default prior implemented in INLA.

### Supplementary Figures

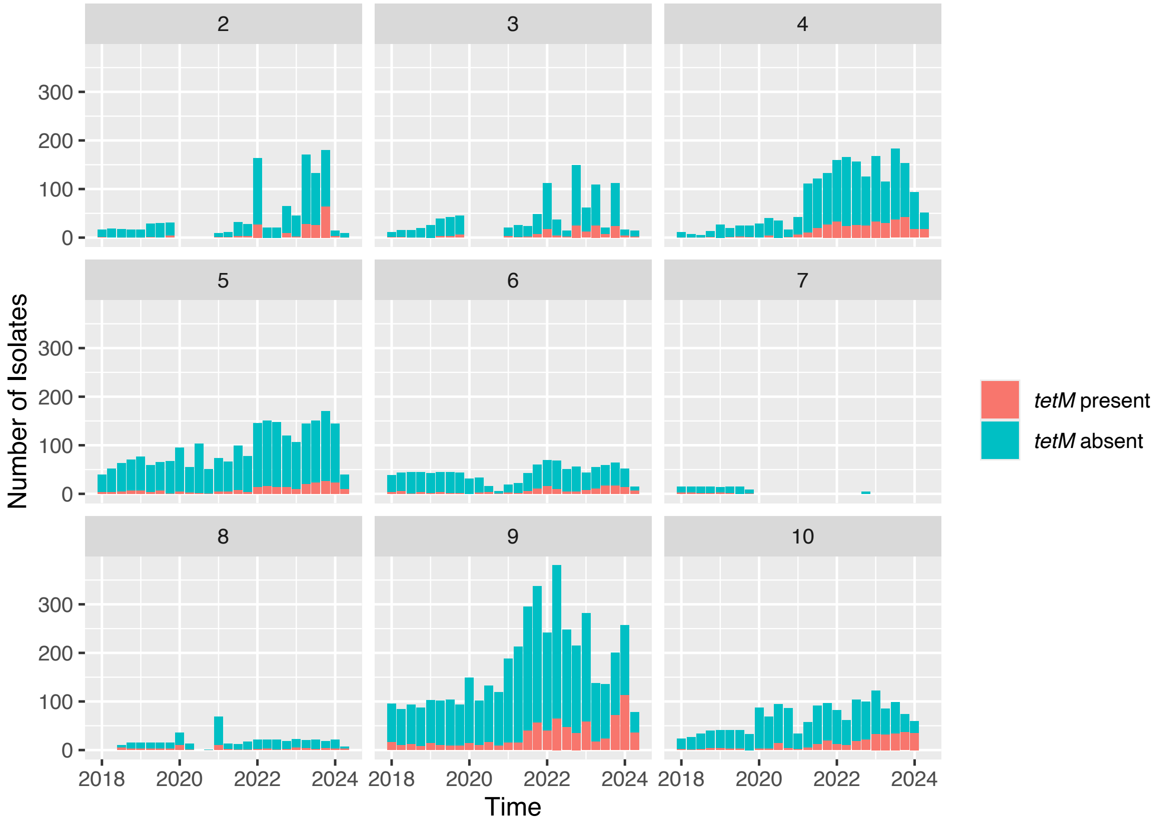


Figure S1: The number of *tetM* positive and *tetM* negative encoding genomes passing quality control thresholds reported each month per HHS region

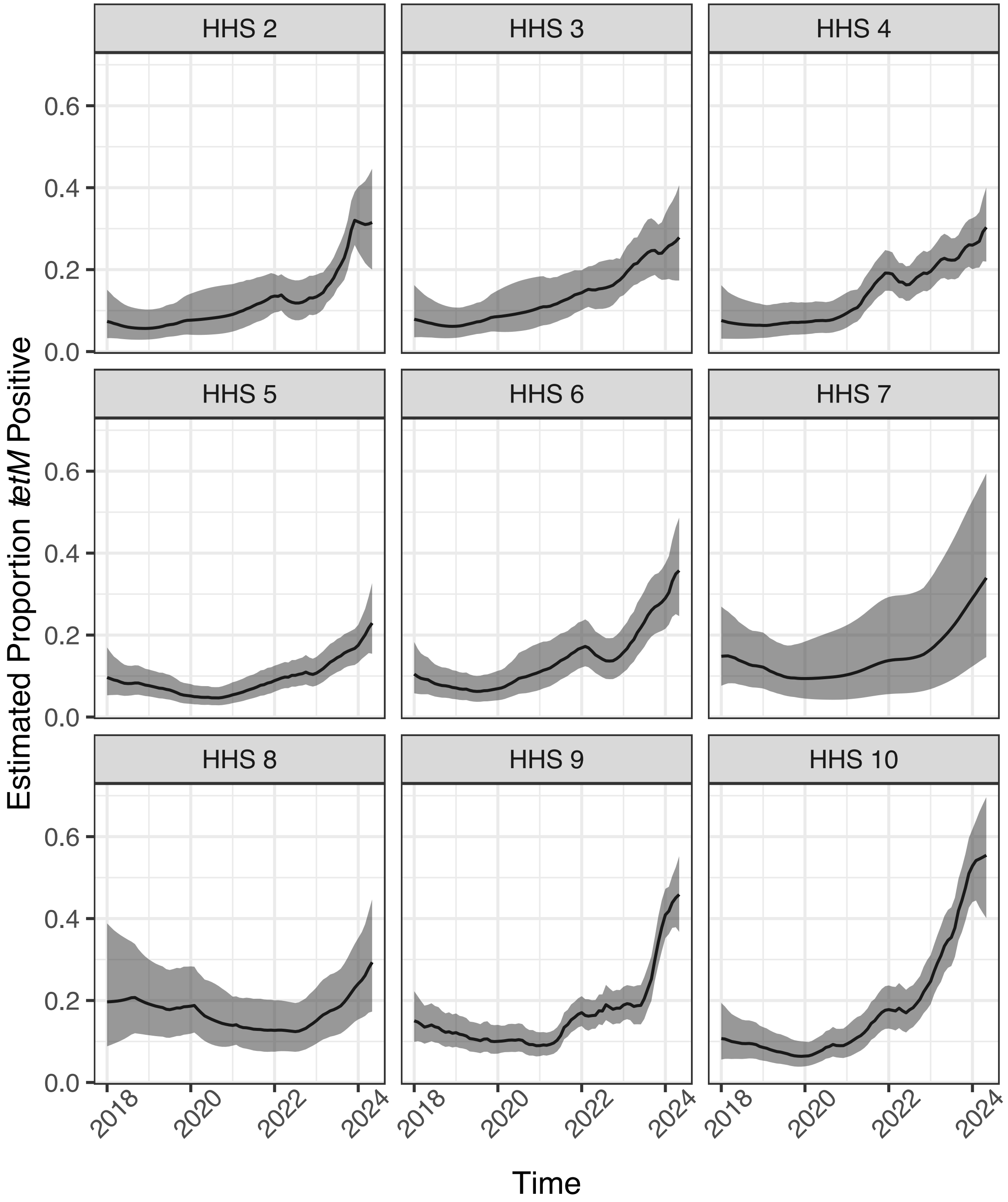


Figure S2: The smoothed estimates of the proportion of *tetM* positive isolates across HHS regions 2-10. Bold black line represents the median estimate. Gray shaded band represents the 95% credible interval around the median.

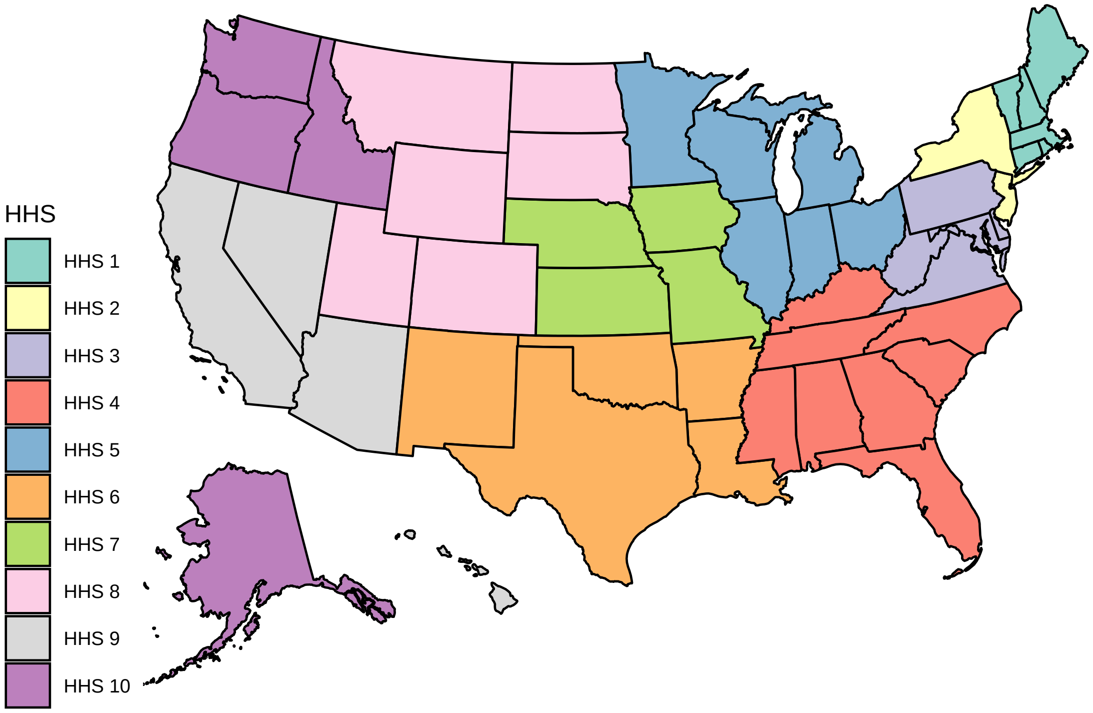


Figure S3: A map of US states with HHS regions indicated.

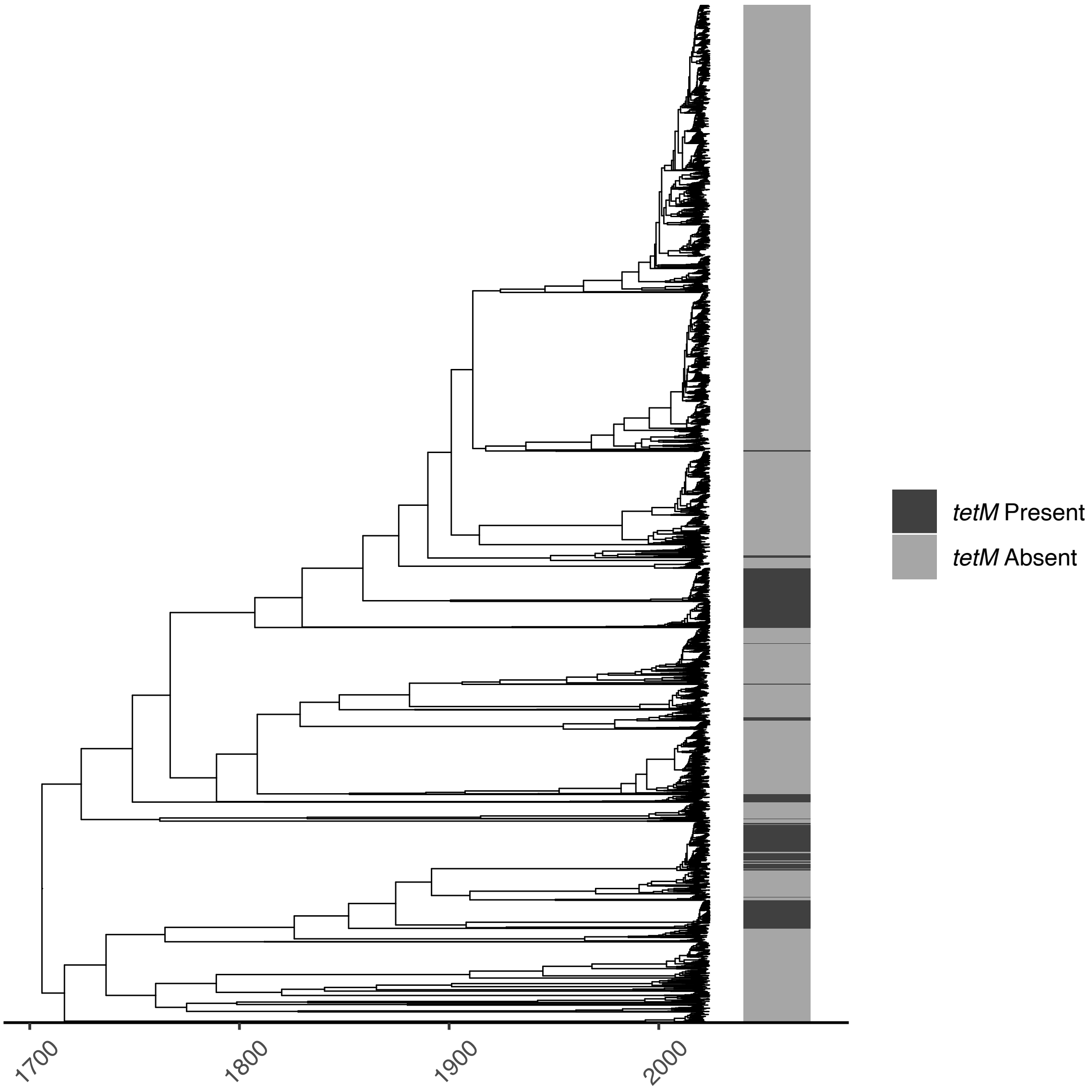


Figure S4: A timed phylogenetic tree of (n=6460) randomly selected isolates collected in the context of CDC *N. gonorrhoeae* surveillance between years 2018-2024. The color bar indicates the presence or absence of *tetM* in the corresponding isolate.
